# A Randomized Sham-Control Trial for Home-Based Transcranial Alternating Current Stimulation (tACS) In Alzheimer’s Disease

**DOI:** 10.1101/2025.04.14.25325464

**Authors:** Lucia Mencarelli, Martina Assogna, Elena Savastano, Ilaria Borghi, Francesca Candeo, Sonia Bonnì, Francesco Di Lorenzo, Roser Sanchez-Todo, Ricardo Salvador, Emiliano Santarnecchi, Giulio Ruffini, Giacomo Koch

## Abstract

1.

**BACKGROUND:** Alzheimer’s disease (AD) is a neurodegenerative disorder for which there is no effective pharmacological treatment. Recent evidence proposes the use of Non-invasive brain stimulation (NIBS) to counteract the typical clinical and neurophysiological degeneration. NIBS protocols require multiple sessions in clinical settings, making long interventions difficult. Therefore, this pilot study aims to evaluate the possibility of moving to a home-based approach.

**METHODS:** We conduct a randomized, double-blinded, sham-controlled, cross-over study of home transcranial alternating current stimulation (tACS) in patients with mild to AD. The intervention consist of daily model-driven personalized home-based tACS delivered for 8 weeks, 5 days per week (40 sessions). tACS is applied daily for 1 hour at 40 Hz on the dorsolateral prefrontal cortex and temporal cortex bilaterally.

The outcomes are assessed at baseline, right after the intervention, and at 2 months follow-up for both conditions (Real and Sham). The primary outcomes measures are the safety and feasibility of the intervention, as well as the changes in the AD Assessment Scale Cognitive Subscale (ADAS-Cog) 13. To assess secondary outcomes, participants undergo extensive neuropsychological assessment. Finally, as exploratory outcomes, functional and structural magnetic resonance imaging (MRI), resting and evoked electroencephalography (EEG and TMS-EEG), as well as blood biomarkers to measure changes in neurodegenerative and neuroinflammatory markers, are collected. The study protocol has been approved by the local institutional review board (IRB).

**RESULTS:** We hypothesize that the tACS treatment is safe and feasible and will result in a slower progression of cognitive decline. Moreover, we wish for a restoring of gamma power as measured by TMS-EEG, as well as a structural preservation and functional network reorganization as compared to sham treatment.

**DISCUSSION:** The final goal is to obtain preliminary data in advance of a larger clinical trial to prove the possibility of treating AD patients at home through tACS.

## 2. Background

Alzheimer’s disease (AD) stands as the foremost cause of dementia, with its prevalence on an upward trajectory. Despite the substantial disease burden and considerable scientific inquiry, therapeutic avenues remain somewhat constrained, with pharmacologic interventions offering only transient stabilization of cognitive functions (1–4). However, recent findings have showcased the potential of innovative non-invasive brain stimulation (NIBS) techniques in enhancing cognition among AD patients (5–9). NIBS enables direct manipulation of brain function in a noninvasive, painless, and safe manner, boasting high spatial precision and commendable time resolution (10– 12).

In AD, cognitive deterioration is intricately associated with the accumulation of protein aggregates in the brain, notably amyloid-β (Aβ) and phosphorylated tau (p-tau), alongside alterations in oscillatory activity (13). The histopathological signatures of AD encompass widespread Aβ plaques and p-tau deposits forming neurofibrillary tangles (14–16). Both Aβ and p-tau are pivotal in the pathogenesis of AD, highlighting the potential clinical significance of interventions capable of safely and effectively reducing their cerebral load. Moreover, a consistent finding in patients with AD is a relative attenuation of the brain’s oscillatory activity in the gamma range (17,18). Gamma oscillations are prominent across multiple brain regions, including the hippocampus (19–21), and are involved in various higher brain functions, including long-term memory, working memory, attention, and sensory processing. The dysregulation of gamma activity has been further linked to pathologic network hyperexcitability in animal models of AD. A recent study found that exogenously induced 40 Hz gamma oscillations reduce Aβ levels and amyloid plaques as well as p-tau levels in a mouse model of AD (22). The authors also showed that in pre-symptomatic AD mice gamma induction prevents subsequent neurodegeneration and behavioral deficits, suggesting that induction of gamma activity may be a powerful new therapeutic approach. Furthermore, recent evidence points toward specific alterations of high-frequency activity within the gamma band resulting from dysfunction of GABAergic parvalbumin+ (PV+) inhibitory interneurons or associated circuits (18,23,24). Reaching this specific neurophysiological substrate requires, among other aspects, the optimization of stimulation solutions able to entrain gamma activity. Modulating interneuron function may improve brain rhythms and cognitive functions in AD and related disorders (18).

Among the array of NIBS techniques available, several studies point out the possibility of using transcranial alternating current stimulation (tACS) at gamma frequency to safely modulate gamma activity in the brain of healthy human individuals (25,26) with corresponding behavioral and cognitive changes both in sensory and high-order cognition domains. More recent studies (5,8,9) concluded that tACS seems to represent a safe and feasible option for gamma induction in AD patients, with preliminary evidence of a possible effect on protein clearance partially mimicking what is observed in animal models, thus increasing blood perfusion (8) and reducing tau burden (27) in temporal regions.

The goal of NIBS is to influence neuronal populations and create favorable conditions for a transition from unhealthy to desired dynamics during or after stimulation. Therefore, we hypothesize that the regulation of the gamma rhythm might improve AD symptoms and pathophysiology. However, a significant constraint of many NIBS protocols is their reliance on multiple sessions to achieve enduring effects, typically conducted in clinical settings necessitating patient travel. This obstacle hampers the delivery of extended interventions, particularly for individuals with neurodegenerative conditions. We thus adopt a home-based approach using tACS because of its ability to entrain gamma oscillations, commonly impaired in AD, and its low cost and potential for home-based application.

We here propose a cross-over home-based tACS pilot study for AD treatment paired with extensive cognitive, physiological, and neuroimaging investigations at three time points to better characterize patients and their response to treatment. The physiological target of treatment is to increase gamma activity in the prefrontal cortex, as this has been associated with cognitive decline in AD. Treatment is multisession since prior transcranial electrical stimulation (tES) work indicates a cumulative effect of each session with stronger therapeutic effects, in line with the underlying Hebbian mechanisms putatively involved in non-invasive brain stimulation.

## 3. Methods

We are conducting a pilot randomized, double-blinded, sham-controlled, cross-over study of model-driven personalized home tACS in patients with mild to moderate AD. The intervention consist of daily model-optimized and individualized 40Hz-tACS delivered on the dorsolateral prefrontal cortex and temporal cortex bilaterally for 8 weeks, 5 days per week (40 sessions, Figure 1). 40Hz-tACS is applied daily for 1 hour. The purpose of this study is to obtain preliminary data in advance of a larger clinical trial designed to test whether repeated, daily sessions of at-home gamma-tACS can lead to a clinically significant improvement in patients with AD. Given the potentially fragile patient population, we propose a pilot study to test feasibility and safety (primary) in this pilot study. The study has been approved by the review board and ethics committee of the Santa Lucia Foundation and it has been registered with the number NCT06826261. All patients or their relatives or legal representatives provide written informed consent.

**Figure 1.**
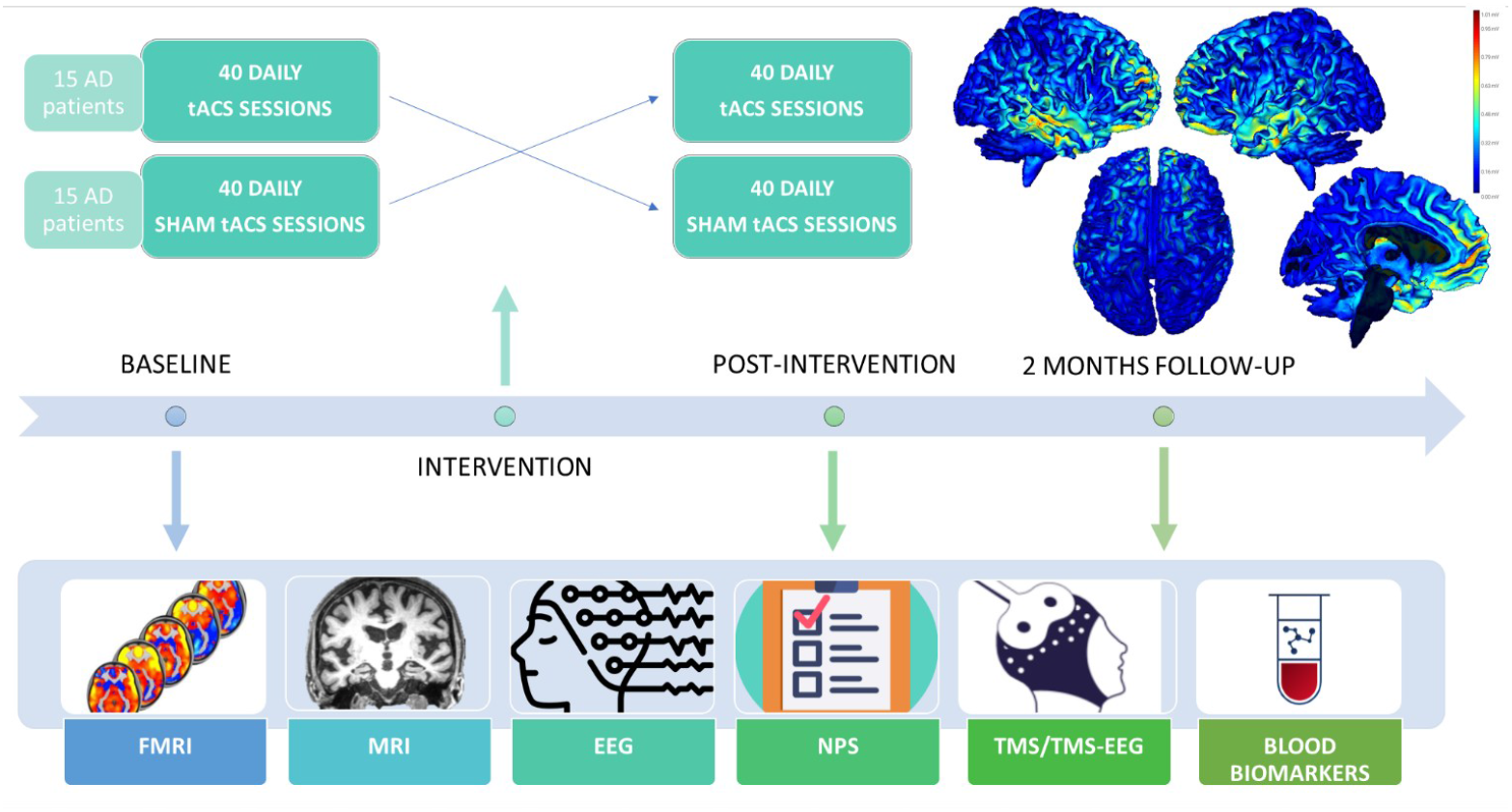
Study design. The patient undergo both the real and sham stimulation in a crossover study. Before and after the treatment, as well as in a 2 months follow up, an extensive assessment is carried. Abbreviations: EEG, electroencephalography; NPS, neuropsychological assessment; TMS, transcranial magnetic stimulation; tACS, transcranial alternating current stimulation; MRI, magnetic resonance imaging; fMRI, functional magnetic resonance imaging.

### 3.1 Study design

This is a cross-over study with Sham and Active tACS conditions (randomized) spaced by a wash-out period (2 months). Neuroimaging, neurophysiological, cognitive data, and blood biomarkers of neurodegeneration are collected at three time points: pre and post-stimulation as well as in a 2-month follow-up (Figure 1). The study is conducted according to the Declaration of Helsinki.

### 3.2 Participants

We enroll 30 mild to moderate AD patients according to the diagnostic criteria International Working Group (IWG) for Alzheimer’s disease (28), based on the following inclusion and exclusion criteria.

#### 3.2.1 Inclusion Criteria

The patients enrolled observe the following inclusion criteria: i) aged between 50 and 85; ii) Clinical Dementia Rating score (CDR) of 0.5-1; iii) Mini-Mental State Examination (MMSE) score of 18-26 at screening indicating mild to moderate Alzheimer’s Disease; iv) treated with acetylcholinesterase inhibitor for at least one month; v) evidence of low β-amyloid and/or elevated phosphorylated Tau protein as detected by lumbar puncture for cerebrospinal fluid biomarkers analysis for diagnostic purposes or Positron Emission Tomography (PET); vi) has an ‘Administrator’; vii) has access to wireless internet (wifi) connection in the location where study treatments is applied.

Moreover, the ‘Administrator’ is examined by the Principal Investigator (PI) to ensure his/her capacity to perform the following in accordance with the requirements of the study protocol for the duration of the study, as explained by the study staff and/or P.I.: i) administer the home-based tES; ii) be accessible to study staff; iii) communicate any safety concerns, potential protocol violations, and any other study-related matters to the study staff or Principal Investigator promptly; and iv) provide the Informed Consent.

#### 3.2.2 Exclusion Criteria

Exclusion criteria include: i) significant neurodegenerative disorder of the central nervous system other than AD; ii) significant intracranial focal or vascular pathology verified by an MRI scan; iii) history of seizures (except febrile seizures in childhood); iv) Diagnostic and Statistical Manual of Mental Disorders, Fourth Edition – Text Revision (DSM IV-TR) criteria met for any of the following (within the specified period): Major depressive disorder (current), Schizophrenia (lifetime), other psychotic disorders, bipolar disorder, or substance (including alcohol) related disorders (within the past 5 years); v) contraindications to MRI (this includes metal implants in the head, pacemaker, cochlear implants, or any other non-removable items if they are contraindications to MR imaging); vi) treatment currently or within 3 months before Baseline with any of the following medications: typical and atypical antipsychotics (i.e., Clozapine, Olanzapine); antiepileptics drugs (i.e., Carbamazepine, Primidone, Pregabalin, Gabapentin); vii) skin lesions on the scalp at the proposed electrode sites; viii) previous surgeries opening the skull leaving skull defects capable of allowing the insertion of a cylinder with a radius greater or equal to 5 mm; ix) any condition that makes the study subject, in the opinion of the investigator, unsuitable for the study.

### 3.3 Home-based tACS Intervention

In our study, the stimulation is applied using the multifocal Starstim device, with current delivered via four Starstim sponge electrodes (circular electrodes with a contact area of 8 cm^2^ embedded in the headcap sized for each patient. All study subjects use the same fixed electrode montage, and the electrode locations are based on the international 10-20 system for EEG.

tACS is applied at gamma frequencies (40 Hz) over the dorsolateral prefrontal cortex and temporal cortex bilaterally (stimulation electrodes: AF3, AF4, T7, T8). These electrode positions are obtained from a group montage optimization using the Stimweaver algorithm (29) performed on a database of biophysical head models of patients with AD, i.e. volume conductor models built from structural head T1w-MRIs that model the passive electrical properties of the primary head tissues. A common montage with only four electrodes is used, to reduce the complexity of the stimulation set-up at home. In order to decrease the variability of the E-field distribution across subjects, the frontal (*±I*_*frontal*_, in electrodes AF3/AF4) and temporal (*±I*_*temporal*_, in electrodes T7/T8) currents is personalized, always assuming a 180° phase-shift between the currents in the electrodes in the opposite hemispheres. This is done using a linear model that predicts the currents based on morphometric characteristics of the patient’s scalp, namely the head perimeter along axial, sagittal, and coronal planes. The full details about the head model database creation, group montage optimization, and creation of the linear regression model are included as supplementary material. The steps of this pipeline (*Neurotwin Xpress* pipeline) are shown in Figure 2.

**Figure 2.**
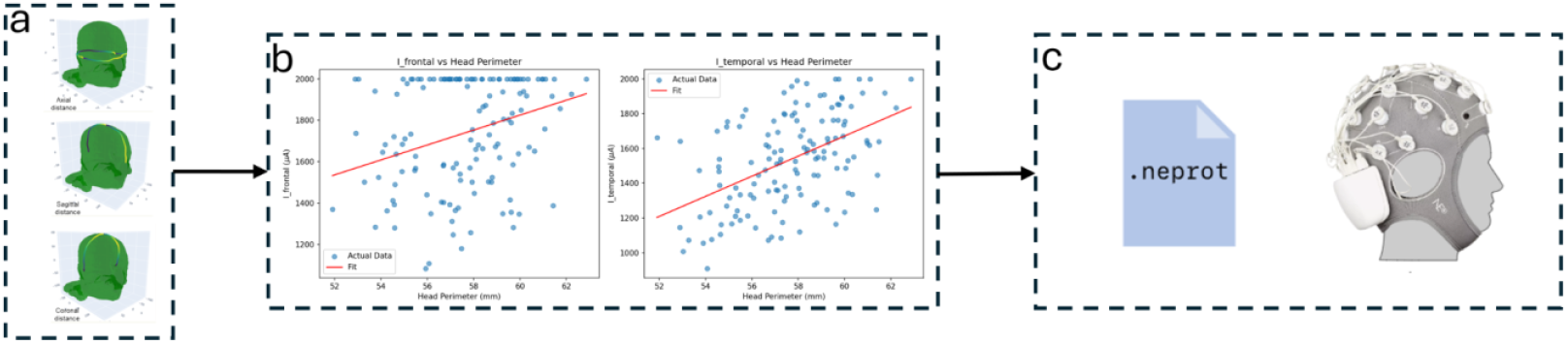
Steps in the *Neurotwin Xpress* personalization pipeline. (a) The perimeters of the head measured along three distinct planes are used as an input to determine the currents. (b) The frontal and temporal currents of the fixed electrodes in the montage are determined via a linear regression model trained on data from a biophysical head models database of AD patients. (c) The currents are then used to generate active or sham protocols (.netprot format uploaded in the NE portal) for tACS stimulation at 40 Hz.

The maximum current for each electrode is constrained to be 2 mA (in absolute value) with a 4 mA max total across all electrodes (sum of currents in all anodes), well below accepted safety limits and considered tolerable in most cases. Prefrontal and temporal electrodes are stimulated in phase (180°). The current intensity is ramped up over the first 30 seconds, then sustained at the stimulation intensity for 60 minutes, then ramped down over the final 30 seconds for the Real condition. The Sham condition rely on the classical 30-second ramp-up/ramp-down protocol.

The subject receive a pre-configured Starstim Home Kit, including the device, tablet, all needed supplies, and training materials. The Starstim Home Kit tablet contains a sequence of simplified instructions and step-by-step touchscreen prompts for the participant/administrator to follow. This process has been designed to be easy to use, even for individuals who are not computer savvy.

The first two daily visits are performed at the memory clinic, then telemedicine session could be set up to further train the administrator in how to properly use the tES Starstim device at home if they request it. Continued support in device use is available to the participants and administrators at any time.

All the home-based stimulation sessions are remotely supervised by the researchers who received information about impedance, tACS progress, answers to final questions, and, potentially, session aborts or interruptions in real-time via email and the NE portal. The Starstim device contain a safety block that only allows a single tACS session to be administered within 24 hours unless otherwise determined by the study staff.

At the end of each daily visit, participants complete a self-report measure to address tolerability.

### 3.4 Outcome measures

#### 3.4.1 Safety and feasilibility

The primary aim of this investigation is to evaluate the feasibility and safety of administering 40 Hz tACS to mild to moderate AD patients using a home-device system. Data about electrode impedance, tACS progression, and session interruptions or terminations, whether voluntary or due to technical issues, are collected by the NE portal. These metrics aid in the assessment of feasibility, including the number of missed sessions. At the same time, any adverse events is documented, and used as the primary safety endpoint.

#### 3.4.2 Neuropsychological and behavioral assessments

Neuropsychological and behavioral assessments are measured by assessing the changes from baseline to the follow-up periods in the following neuropsychological tests: i) The Alzheimer’s Disease Assessment Scale 13 (ADAS-Cog _13_) (30); ii) Clinical Dementia Rating scale (C.D.R.) sum of boxes (31); iii) AD Cooperative Study - Activities of Daily Living (ADCS-ADL) (32); iv) Mini-Mental State Examination (MMSE) (33); v) Frontal Assessment Battery (F.A.B.) (34); vi) Neuropsychiatric Inventory (NPI) (35); vii) Quality of Life Enjoyment and Satisfaction Questionnaire Short Form (Q-LES-Q-SF) (36); viii) Apathy Motivation Index (37); ix) Apathy Motivation Index – caregiver version (38).

#### 3.4.3 Exploratory Endpoints

The following neurophysiological and imaging measures are employed to conduct an exploratory assessment of the effects of 40Hz-tACS on cortical activity, focusing on alterations from baseline to the follow-up periods.

##### MRI assessment

To evaluate the potential modifications induced by the 40 Hz tACS administration on structural and functional imaging, the MRI assessment is collected at baseline, right after the intervention, and at 2 months follow-up for both conditions (Real and Sham). The MRI imaging is conducted on a 3T Siemens Magnetom Prisma scanner with a 64-channel head-neck coil at the Neuroimaging department of IRCCS Santa Lucia Foundation (Rome, Italy). The subjects are instructed to maintain their eyes open fixing a cross, not to focus their thoughts on any particular topic, and not to cross their arms or legs.

##### EEG

Resting state EEG is obtained by using 64 channels of TMS-compatible EEG equipment (BrainAmp 32MRplus, BrainProducts GmbH, Munich, Germany) with the eyes open for five minutes. TMS-compatible Ag/AgCl pellet electrodes are mounted on an elastic cap, while additional electrodes are used as ground and reference. The ground electrode are positioned in AFz, while the reference one is positioned on the nose tip. The EEG signal is band-pass filtered at 0.1–1000 Hz and digitized at a sampling rate of 5 kHz. Skin/electrode impedance are maintained below 5 kΩ.

##### TMS procedures

To evaluate the neurophysiological modifications induced by the 40 Hz tACS, we use paired-pulse (SAI) and long-term potentiation (LTP) TMS protocols (39,40). These protocols allow us to evaluate in vivo the activity of different intracortical circuits in the primary motor cortex. For these measurements, motor evoked potentials (MEPs) are recorded from the right first dorsal interosseous muscle using Ag/AgCl surface cup electrodes. A monophasic Magstim 200 device is used to define the motor hotspot and to assess MEP size using a standard 70-mm figure-of-eight– shaped coil. The motor hotspot is defined as the location where the TMS pulse produced the largest MEP size at 120% of the resting motor threshold (RMT) in the target muscle.

##### TMS-EEG

Participants undergo TMS-EEG protocols before, right after the intervention, and at 2 months follow-up for both conditions (Real and Sham) to investigate changes in cortical reactivity, connectivity, and oscillatory evoked activity. We use 64-channel TMS-compatible EEG equipment (BrainAmp 32MRplus, BrainProducts GmbH, Munich, Germany) to collect data. A masking noise system is used to reduce auditory evoked potential artifacts. The stimulation take place over multiple brain regions in the prefrontal and parietal lobes. TMS intensity is set firstly by computing the RMT. Since the coil-to-cortex distance directly influences the magnitude of magnetic stimulation, for each patient, we subsequently calculate a distance-adjusted RMT (AdjRMT)(41). Procedures for neuronavigation, TMS delivery, and EEG preprocessing/analysis will reflect validated procedures previously published by our group (42,42,43).

##### Blood biomarkers

Blood biomarkers are collected in order to analyze changes in neurodegeneration biomarkers (Neurofilament light NFL, Beta-amyloid, p-tau, total-tau) at baseline, at the end of the intervention, and at 2 months follow-up. We measure the concentrations of NFL in ethylenediaminetetraacetic acid–treated plasma samples using the single-molecule array (SIMOA) immunoassay (44). Measurements are performed on coded samples. All laboratory personnel remain blinded to treatment allocation and diagnosis and have no access to clinical data. We also collect Plasma Aβ42, Aβ40, and t-tau concentrations simultaneously using the SIMOA-HD1 platform (SIMOA; Quanterix, Billerica, MA, USA), which employs an automated SIMOA principle. Briefly, Aβ42, Aβ40, and t-tau levels are measured using a multiplex array (Neurology 3-Plex A Advantage Kit, N3PA). The samples are measured using a two-step immunoassay (45).

#### 3.5 Statistical Analysis

All the outcome measures are analyzed by calculating the change from baseline after 8 weeks of treatment and 2 months follow-up using a mixed-model repeated-measures analysis. The null hypothesis is that the difference between the two conditions (Real and Sham) right after the intervention is equal to zero. All the analyses involve an Analysis of Covariance (ANCOVA) model incorporating the baseline cognitive/clinical scores as covariates. Furthermore, each neurophysiological parameter is used in univariate and multivariable (considering clinical/demographical/APOE genotype as possible confounding factors) regression analysis to assess the potential predictive value of each parameter in response to 40 Hz tACS treatment. All variables will be summarized using descriptive statistics, i.e., number (%) of patients for categorical variables and mean, SD (standard deviation), median, minimum, and maximum for continuous variables.

Correction for multiple comparisons will be used where appropriate. For all statistical analyses, a p-value of <0.05 will be considered to be significant.

## 4. Discussion and expected results

Given the absence of effective treatments for Alzheimer’s disease (AD), there is a pressing need for the development of new interventional strategies aimed at attenuating disease progression. Drawing upon the collective experience garnered from numerous clinical trials using NIBS on AD (5– 9,27,41) which have documented promising evidence regarding enhancements in cognitive performance, activities of daily living, and brain oscillations, the proposed protocol of 40 sessions of daily 1-hour home-based 40 Hz tACS may represent a promising therapeutic avenue for AD patients. Previous studies have typically employed multiple sessions of rTMS or tACS within clinical settings (8,27,41). While these studies have yielded encouraging results, the burden on patients and caregivers has been considerable. Consequently, transitioning to home-based stimulation may offer greater accessibility to numerous patients. Although rTMS may have a more substantial clinical impact compared to electrical stimulation techniques (46), it necessitates expensive equipment and must be administered by specialized personnel in hospital settings. In contrast, our decision to utilize tACS in this study stems from its affordability, ease of use, suitability for administration by non-specialists, and potential for home-based application, thus reducing distress and costs associated with institutional-based care and increasing patients’ compliance.

The primary goal of our study is to establish the safety and feasibility of administering 40 Hz tACS for 1 hour per day over 40 sessions in patients diagnosed with AD. We aim to achieve this objective by ensuring that no serious adverse events occur during the intervention. Furthermore, we assess feasibility based on whether the therapy is successfully delivered for at least 75% of the total programmed time. A session will be considered missed if less than 75% of the scheduled duration is completed by the subject.

Considering the cognitive implications, based on prior reports on the impact of γ-tACS on AD pathophysiology (17,18,47), we anticipate that real 40 Hz tACS will result in stabilization of memory and executive function scores compared to sham condition. These cognitive enhancements could potentially lead to indirect improvements in other health-related outcomes, including quality of life and independence in daily living activities, consequently alleviating caregiver burden.

From a biological standpoint, in this study, we anticipate that multiple sessions of 40 Hz tACS result in increased power of fast brain oscillations, accompanied by decreased power of slower oscillations. Moreover, we hypothesize that this effect may extend beyond the targeted regions to involve other brain areas that are structurally and functionally connected. Furthermore, we assume that functional connectivity and structural volume would be modulated by the 40 Hz real tACS but not by the Sham condition. At the same time, cortical reactivity, oscillatory patterns, and connectivity at high temporal resolution registered through TMS-EEG would be helpful in evaluating the efficacy of 40 Hz tACS. Additionally, blood biomarkers would unveil the potential role of tACS in neurodegeneration and neuroinflammation; and TMS protocols would measure the synaptic transmission deficit before and after the treatment. Furthermore, to explore individual differences in response to 40 Hz tACS, we collect EEG data at each time point. This would enable us to identify new markers that characterize the brain’s responsiveness to tACS in AD. Subsequently, we aim to retrospectively identify “responders” and characterize the individual trajectories of response to 40 Hz tACS. Although the intensity of stimulation is tailored to individual head measurements (see supplementary material for more information), we opted for a frontotemporal standard montage that could be applied to a broader population of AD patients if successful. This approach will enable us to assess the correlation between behavioral and clinical effects and the accuracy of the template stimulation solution for each participant. This analysis has the potential to elucidate the variability between responders and non-responders, thereby informing future trials regarding the necessity for personalized montages.

Finally, the two-month follow-up period provides an opportunity to evaluate the sustained efficacy and durability of the 40 Hz-tACS intervention over an extended timeframe, offering valuable insights into its long-term effects on cognitive function and overall outcomes. In addition, digital twin models of each participant (*neurotwins*) are created through data assimilation (anatomical MRI, tractography, fMRI-BOLD and ASL, EEG, TMS-EEG) to evaluate pre/post changes through model parameters. This allow us to verify mechanistic hypotheses of the intervention, in particular, the recovery of fast oscillatory circuits from tACS entrainment as reflected in local connectivity parameters of PV+ interneurons.

In conclusion, the urgency for effective treatments for AD patients necessitates innovative interventional strategies to mitigate disease progression. Our proposed protocol of home-based 40 Hz-tACS treatments for AD patients, either standalone or combined with other therapies, could revolutionize treatment options and thus represent a crucial step in unveiling tACS mechanisms for AD intervention, informing future trial designs.

## Data Availability

All data produced in the present work are contained in the manuscript

## Acknowledgements

The clinical trial is supported by the European Union’s Horizon 2020 research and innovation program under grant agreement No 101017716.

## Authors’ contributions

Conceptualization: GK, LM, MA, GR; Funding acquisition: GK, GR, ESan; Data curation: LM, ESav; Statistical modeling and design: GK, GR; Data collection: LM, ESav, MA, FC, FDL, IB, SB; Project administration: LM, ESav; Brain Stimulation solution: RST, RS, GR; Supervision: GK, GR, ESan; Manuscript Draft: LM, RS. All authors have substantively revised the work and approved the submitted version.

